# The blurred threshold of AI-use disclosure: International journal editors’ expectations of sufficiency and necessity

**DOI:** 10.1101/2025.07.17.25331725

**Authors:** L. Lingard, E. Driessen, K. Oswald

## Abstract

**Purpose:** Generative AI is a powerful resource for health professions education (HPE) researchers publishing their work. However, questions remain about its use and guidance about disclosure is inconsistent. This situation is both confusing and potentially perilous for researchers, who risk their reputations if they disclose AI use inappropriately. This study explores HPE and general medical journal editors’ experiences and expectations of AI-use disclosure, in order to assist journals to clarify expectations and authors to satisfy them.

**Methods:** In this descriptive qualitative study, journal editors were interviewed between January 6, 2025, and May 7, 2025 using an online Zoom platform. Eligible participants were identified through journal webpages and snowball sampling. A purposive sampling strategy prioritized HPE research journals and included a limited sample of general medical research journals to explore transferability. Data collection and thematic analysis proceeded iteratively.

**Results:** Eighteen participants, including 9 chief editors and 9 associate/deputy editors were interviewed. Fourteen participants worked in HPE journals, four in general medical journals. The analysis revealed 4 key themes: 1) the basics of disclosure, made up of content and location expectations shared by participants; 2) the sufficiency threshold, regarding how much detail to include; 3) the necessity threshold, regarding which circumstances require disclosure; and 4) the factors blurring these two thresholds, which included the speed of change, the co-construction of disclosure standards, and the uneasy fit of scientific principles such as reproducibility and transparency with the AI-use context.

**Conclusions:** While editors shared basic disclosure expectations, they also provided insight into blurred thresholds of sufficiency and necessity that complicate disclosure. By attending to these thresholds and the factors blurring them, and by using these insights to apply recent AI-use and disclosure frameworks, journals can develop enhance their guidelines, which will assist authors in HPE in navigating the shifting norms of AI-use disclosure.

## Introduction

GenAI offers a powerful resource for health professions education (HPE) researchers publishing their work^1,2,3^. However, questions remain about appropriate use^4^ and published guidance is inconsistent^5^. This situation is confusing and potentially perilous: researchers risk their reputations by disclosing inappropriately.

The initial debate about *whether* AI should be used to help write manuscripts^6,7,8^ was short lived; surveys^9^ and database analyses^10,11^ demonstrate that this is emerging practice. Although we agree that AI cannot be an author, debate persists regarding effective^12^ and acceptable^13^ uses, and guidelines often lack specifics^14^. Add a growing concern about undisclosed AI use^15,16^, and it is clear that researchers need more clarity.^9^

Although AI-use disclosure is required for authors in and outside of health professions education (HPE)^17,18,19^, guidance varies. It also evolves over time,^5,20,21^ evidenced by the ICJME’s shift from general recommendations to specific guidance about what and where to disclose.^22,23^ Health professions education journals have followed ICJME’s evolution and require human oversight^24^ and greater transparency from authors.^25^ Some general medical journals have shifted from discouraging AI content in 2023^26^ to anticipating it in 2024.^27^ Such evolution makes sense as AI technology advances. However, it can also create confusion. Researchers disagree^28,29^ and can’t find answers to straightforward questions: when is disclosure mandatory?^30^, which uses should be disclosed?^31,27^, what details are required?^32^, and how will it impact peer review?^33^

Missing from the literature is empirical study of HPE’s journal editors’ perceptions.^5^ Editors have a unique perspective on and significant responsibility for the development of disclosure standards. Therefore, this study asks: *What are journal editors’ expectations and experiences of AI-use disclosure statements in submitted research manuscripts*?

## Methods

We used descriptive qualitative methodology^34^ situated within an interpretivist research paradigm, and followed the SRQR conventions for reporting qualitative research.^35^

### Participants

We purposefully sampled for experienced editors of HPE journals, informed by the Medical Education Journal List 24 (MEJ-24).^36^ Because HPE journals are situated within the medical field, we also included editors from two major families of general medical research journals to explore transferability of the work. Eligible participants were identified through journal webpages and snowball sampling. We determined thematic sufficiency^37^ of the sample with reference to the principle of information power which we sought to maximize through targeted sampling of rich informants and techniques to enhance interview quality.^38^ Participants were invited via email and we prioritized editorial experience in our recruitment.

Data Collection: We conducted individual, semi-structured Zoom interviews lasting 30-50 minutes. Interviews took place between January 6, 2025, and May 7, 2025. Questions asked editors’ expectations regarding disclosures and their journals’ experience of handling them (Supplement 1). Most participants brought anonymized examples of submitted AI-use disclosure statements to discuss.

Data Analysis: Data collection and analysis proceeded iteratively. Using thematic analysis procedures^39,40^, all authors read the first five transcripts and discussed preliminary codes. LL and KO independently identified inductive codes for the next eight transcripts, which were discussed by all authors for consistency, with attention to discrepancies requiring exploration in later interviews. KO re-analyzed all transcripts using the final coding structure, manually extracting all passages relevant to each code. LL and ED reviewed this coding and identified themes. KO re-analyzed all transcripts for themes, preparing analytic memos for discussion. Trustworthiness was enhanced through an audit trail and researcher reflexivity41.

The authors are experienced scientific editors and authors (ED, LL) in HPE and a graduate student (KO) in information and media studies. Their experiences and assumptions about AI-use disclosure were explicitly discussed and managed through techniques such as non-leading interview questions and discrepant case analysis.

This study received institutional ethics approval from Western University’s Non-Medical Research Ethics Board (ID#125269).

## Results

Eighteen participants were interviewed (Table 1); three declined. Nine of the MEJ-24 journals were included, and four general medical journals from two prominent medical journal families. For five journals, we interviewed both the Editor-in-Chief and an associate editor they identified as particularly experienced with AI disclosures. Fourteen participants brought anonymized examples to discuss. All general medical journal editors and all but one HPE editor reported seeing disclosures in submitted manuscripts in the preceding months. Overall, HPE editors conveyed more limited experience with disclosures than general medical editors.

**Table 1:** Participant Demographics

| **Demographic** | **# (%) participants**<br>**N= 18** |
| --- | --- |
| **Journal type** |  |
| Health professions education journal | 14 (77) |
| General medical journal | 4 (22) |
| **Role** |  |
| Editor in Chief | 9 (50) |
| Deputy/Associate editor | 9 (50) |
| **Participant Country** |  |
| Australia | 1 (.05) |
| Canada | 4 (22) |
| Netherlands | 1 (.05) |
| Oman | 1 (.05) |
| Singapore | 1 (.05) |
| UK | 3 (.05) |
| USA | 7 (38) |
| **Gender** |  |
| Female | 10 (55) |
| Male | 8 (44) |
| **Years of editor experience** |  |
| <5 | 1 (.05) |
| >10 | 17 (94) |

Four themes were identified across the dataset: 1) the basics of disclosure, 2) the sufficiency threshold, 3) the necessity threshold and 4) blurred thresholds.

### The basics of disclosure

Participants shared a sense of the basics for reporting AI-use (Figure 1). Most expected authors to name the tool, specify the task it was used for, and include a statement attesting responsibility for the material. They noted positive features in disclosures they brought to discuss:

**Figure 1.**
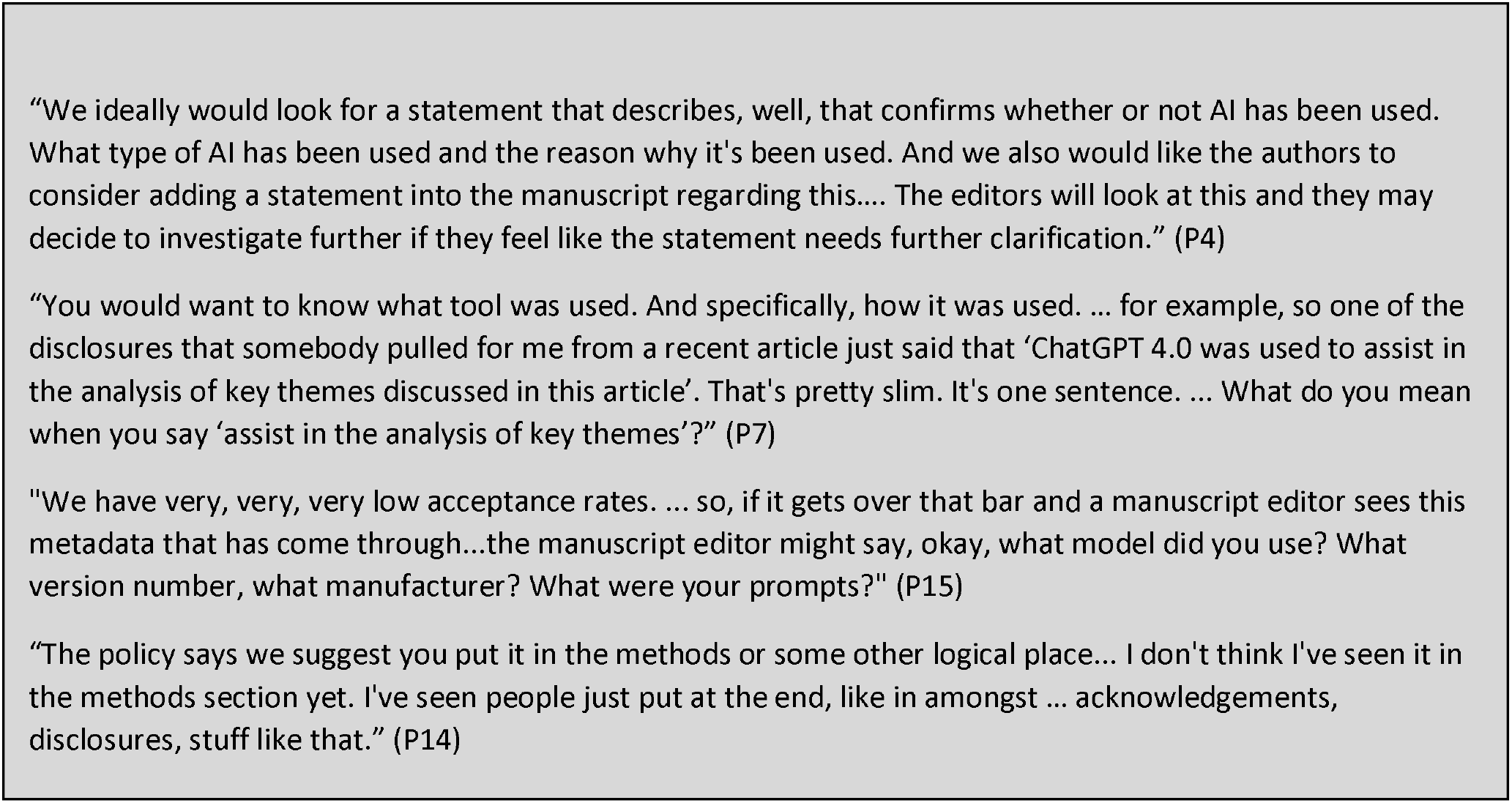
Additional quotations for The Basics of Disclosure

> This author acknowledges that “specific generative AI tools have been used during the writing of this article, was used to suggest paraphrasing of author-written text, and to provide ideas for the structure of the introduction and conclusion sections. Not all of these suggestions or ideas were used, and all AI-generated content was reviewed and adapted by the author prior to submission.” So, now, that’s a nice one. (P17)

In editors’ experience, however, these basics were often missing or mis-placed; disclosures were “a little messy” (P12); and practices were shifting over time: “The disclosures we received in 2023 and 2024 were a lot more detailed than the disclosures that we have received more recently. …what I see now are very short disclosures -- ‘we used AI tools to improve the grammar or flow’-- or whatever” (P13). There was shared interest in standardized templates, but concerns about possible confusion given “a cottage industry now of reporting guidance guidelines for use of AI in research” (P15).

### The sufficiency threshold

Beyond these basics, expectations diverged regarding how much detail was considered sufficient (Figure 2). Some editors wanted authors to “make sure that its fully transparent” (P4), while others preferred disclosure to “scale” (P8) with AI’s impact on the content:

**Figure 2.**
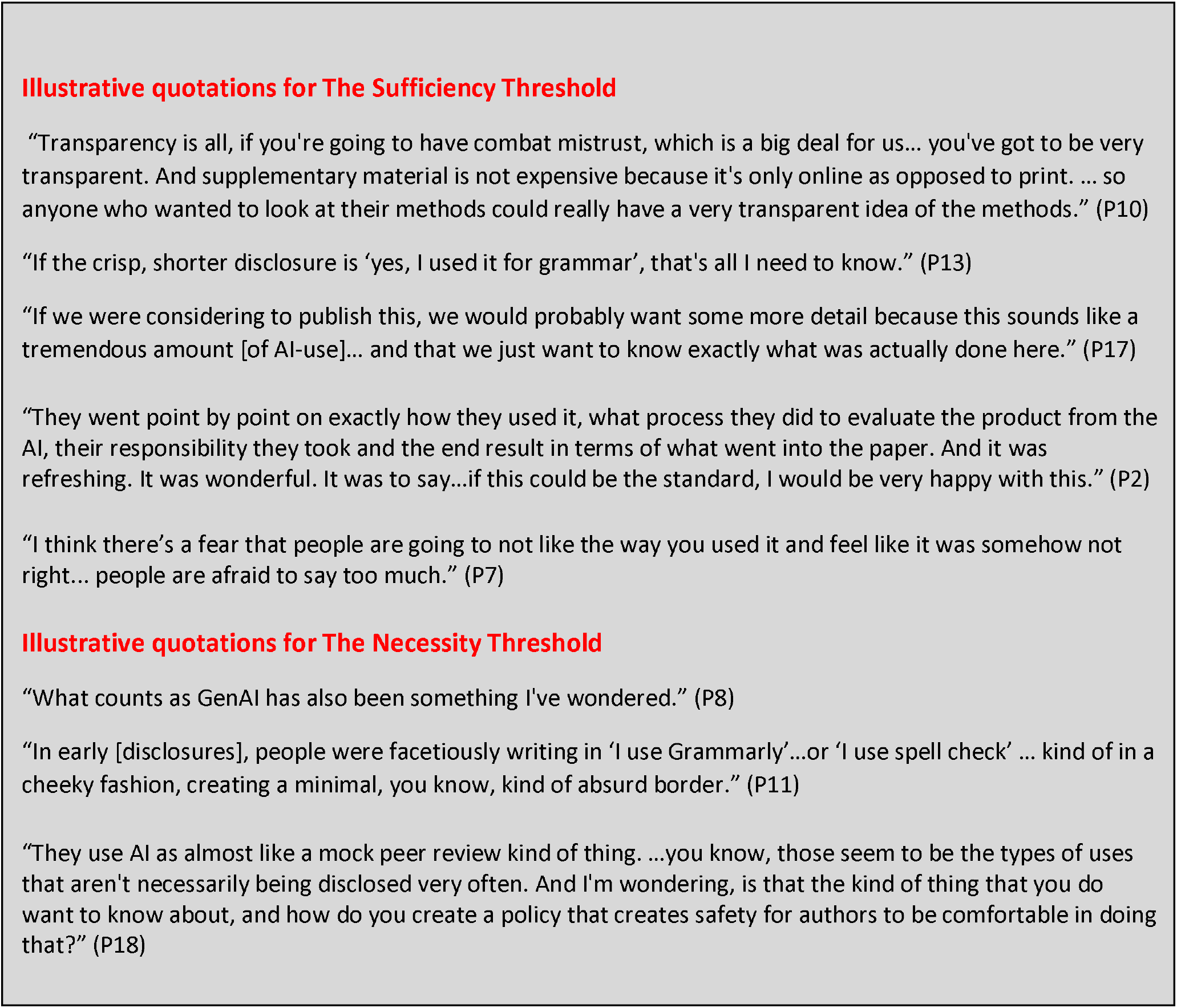
Additional Threshold Illustrations

> I wouldn’t care what they put into their prompt if it just said something like, ‘I wanted a summary abstract’ … but if it was something along the lines of ‘we got the AI tool to do a first draft of our discussion’, I might get a little bit more interested into exactly what they put into that prompt. (P14)

Some suggested supplementary files for prompts or chatlogs, but responses varied regarding when such detail mattered. Editors noted that sufficiently detailed disclosures build trust, while vague ones raise concern. However, participants viewed the potential impact of details differently, recognizing that “there’s a tension between, you know, we say we want you to be transparent, but if we’re transparent you’re going to punish us somehow” (P5).

### The necessity threshold

Participants discussed which uses necessitate disclosure, and which do not (Figure 2). Using AI for spelling or grammar was not perceived to warrant disclosure because such tools have “been around forever” (P10), but editors also acknowledged that “that pushes it back to the author to say, well, what’s AI?” (P2). All editors perceived that substantial use must be disclosed, but they defined this variably. Some put the threshold at “intellectual” (P4) work, but the distinction between superficial and substantive use was acknowledged to be “a very gray line” (P8): “Okay where’s the threshold? I wonder as a scholar, if people just don’t know. That’s one of the reasons that they’re not disclosing, because they’re just not really sure what’s okay and what’s going to get editors and peer reviewers a bit squinty-eyed” (P8).

### Blurred thresholds

Participants discussed three factors blurring these thresholds: the speed of change, the philosophy of co-construction, and the uneasy fit of some scientific principles (Figure 3).

**Figure 3.**
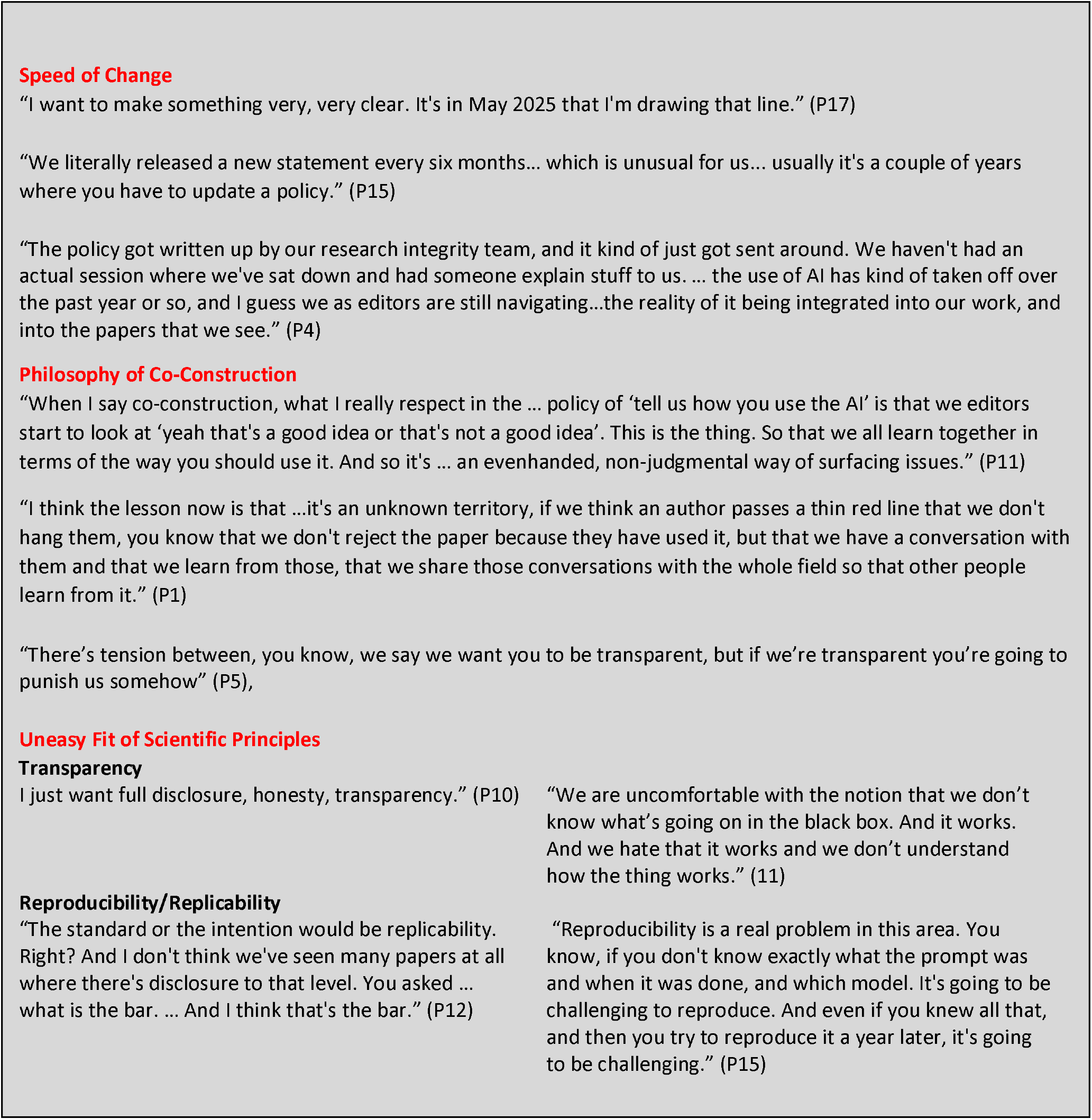
Additional quotations for Blurred Thresholds

Participants reasoned that journal policies remained vague because the speed of change would make any policy outdated: one described the situation as “a very fast-moving space… that we’re going to have to constantly revisit” (P16). Online submission architecture was “a moving target” (P6). HPE editors in particular reflected that “we’re early days… six months ago, sort of we didn’t have a policy like a lot of places” (P7); there was a perception that “the whole field has more questions than answers” (P1). Some described editorial experiences as limited: “it’s very rare still for me to see these disclosures” (P6). And there was a perception that editors “only know philosophically what they think a good one should look like, because they actually haven’t seen enough of them” (P9).

Editors characterized the scientific field as co-constructing an understanding of AI-use disclosure. But participants recognized that co-construction creates uncertainty. Some mentioned the power differential in publishing, recognizing that disclosure statements are viewed as how we “police this situation” (P5) and authors are “worried that [disclosure’s] going to negatively impact their article” (P7). All editors explicitly noted their journal’s non-punitive stance: “the intention is not to punish authors” but to “develop our thinking and or moralities around this topic” (P1). However, there was also a sense that “it’s a bit of a liminal space… in between two worlds we are now. We are also a little bit lost, I think” (P16). Given this context, disclosure was perceived to require courage: “It’s like we’re all using this and who’s going to be the one to step up and be that guinea pig, you know, somebody that’s willing to have those conversations without being redlined or not published in these journals. Right. That’s really hard to do” (P1). Editors also worried that co-construction of disclosure standards could be complicated by inconsistencies between policy and practice: “It says right in there: ‘checking this box will not affect the decision about your article’. That being said, I’ve seen conversations in the background where people said,’ oh, they used AI, we need to discuss this’. So, I think we’re talking out of both sides of our mouth” (P7).

Disclosure thresholds were also complicated by the application of incompatible scientific principles, particularly reproducibility and transparency. Many participants advocated disclosure to ensure research would “be reproducible” (P12), and wanted sufficient information “to interpret, to appraise, to possibly replicate the work” (P6). Others explicitly dismissed reproducibility as incompatible with AI: “you can put the same prompt into it twice in a row and get different response – there’s nothing reproducible about your interactions with genAI” (P11). Transparency was also recurrently invoked as a criterion for robust disclosure: “the most transparent you can be is providing the actual prompts, almost like a script of what you did. That’s like, ‘here’s what I did. Go for it. Look at it all you want.’” (P7) However, other editors questioned how transparent a disclosure could be, given the “black box” (P17) of AI and users’ poor understanding of how it produces outputs.

Table 2 summarizes insights from our thematic results into disclosure guidance for authors. ChatGPT 4o was used to prepare a draft, which we edited significantly; we take responsibility for this version.

**Table 2.** AI Disclosure Guidance for Authors based on Editor Expectations

| Disclosure Basics | Complexities of Sufficiency and Necessity |
| --- | --- |
| **State how you used AI.**<br>Example: “We used ChatGPT 4o (May 2024 version) to shorten our abstract and strengthen our study limitations.” | **Distinguish between technical & intellectual use.** Intellectual uses are of more concern to editors and require more explanation. Example: “We used ChatGPT 4o (May 2024 version) to help brainstorm counterarguments in the Discussion. We prompted counterarguments by discipline (e.g., psychology) and research paradigm (e.g., positivism).” |
| **Be specific: tool, tasks, location.**<br>Include at minimum: the tool name, version or date (if known), the tasks performed, and the manuscript section(s) affected. | **Be strategic about the level of detail.**<br>More detail is not always better or feasible. Scale down details for technical use (e.g., editing grammar); scale up for more intellectual use (e.g., creating interview prompts). Editors may request more details as the manuscript nears acceptance. |
| **Add a responsibility statement.**<br>Attest that you are accountable for the content. Example: “All AI-assisted content was reviewed and verified by the authors, who take full responsibility for its accuracy and integrity.” | **Indicate *how* you verified content.**<br>Verification is a process, not a promise. Be transparent about how you verified. Example: “We verified accuracy of AI-produced text by conducting our own PubMed search and reviewing key sources. Where we found discrepancies or oversimplifications, we edited the AI text as needed.” Consider what aspects of AI-use might not be transparent or verifiable. |
| **Put the disclosure in your manuscript.**<br>Editors prefer this in the Methods or other section as relevant, rather than in Acknowledgements. | **Aim for credibility, not necessarily reproducibility.**<br>Share your thinking process; acknowledge inconsistencies in AI output; describe how you iterated prompts to achieve better results. If you complete a checkbox or text box during online submission, ensure coherence with manuscript disclosure text. |
| **When in doubt, disclose.**<br>If the AI use shaped ideas, text structure, or scholarly content—not just grammar—editors generally expect disclosure. | **Don’t avoid disclosure in fear of punitive action.**<br>Journals and authors are co-constructing the norms of disclosure, learning together in a fast-changing situation as AI technology and literacy develop. Disclose in this spirit. |

## Discussion

Given evidence of nondisclosure,^10,42^ clarity of expectations is critical.^43^ Amid a sea of policies^23,44^ and guidelines^45,46^, researchers are divided on^26^, uncertain about^47^, and afraid of^48,49^ AI-use disclosure. In this first study to bring HPE and general medical journal editors’ perceptions to bear on this situation, participants shared basic disclosure expectations and provided insight into blurred thresholds that complicate these basics.

Understanding that disclosure thresholds are blurred, even for editors, can be empowering for authors. The sufficiency threshold flags the challenge of anticipating whether details will be positively or suspiciously received. Some editors advocated maximum detail; others preferred disclosures scaled for AI-use. Timing matters: early advice to include prompts and chatlogs^50^ is changing. The necessity threshold alerts authors to the challenge of judging which circumstances require disclosure. With disclosure still relatively uncommon according to HPE participants, this threshold may be hard for authors to discern. Further, if the HPE research community understands AI is being used but rarely sees disclosure^10,39^, this may encourage assumptions that disclosure is unnecessary.

We identified three factors influencing blurred thresholds. Below we discuss how they could help journals to clarify disclosure expectations and authors to apply them.

The speed of change makes AI-use disclosure a fluid environment. All participants noted this fluidity; many cited it as justification for minimal disclosure guidance. However, a fluid environment requires more, not less, author guidance. With a ‘cottage industry’ of frameworks^51,52,42^, authors need direction. Editors need it to, particularly given HPE editors’ reports of limited experience with AI-use disclosure to date. Some of the emerging resources could help HPE journals to clarify their positions and educate editorial board members; e.g., a recent framework that distinguishes mandatory, optional, and unnecessary disclosure^5^ could stimulate productive discussion and explicit setting of necessity thresholds that are contextually relevant for HPE and temporally appropriate as AI continues to evolve.

Co-construction also blurs the necessity and sufficiency thresholds. Editors expected that authors, by declaring AI-use details, would help journals gradually articulate the dimensions of appropriate AI-use and disclosure. Editors understood that co-construction required authors to take risks, although they described a non-punitive approach to insufficient or absent disclosures. Co-construction might be enhanced in the HPE field – and risks mitigated – by drawing on established publishing norms. For example, journals might use the Artificial Intelligence Disclosure framework^53^ that adapts the CRediT taxonomy^54^ of contributor roles to AI-use to create more precise guidelines for authors. Because it taps into our field’s familiarity with the expectations for authorship contribution, this framework could prove a helpful resource while we co-construct our disclosure norms in a fluid environment.

Finally, our results suggest the need for critical discussion of how longstanding scientific principles fit with AI. While reproducibility and transparency feature prominently in our dataset and in published guidelines^43^ for AI-use and reporting, the inconsistency of AI responses and the black box nature of AI raises questions about their fit. We are not suggesting that journals discard traditional scientific principles, which may be more, not less, important with the rise of AI-use.^24^ However, we do advocate explicit and critical discussion of how these principles shape disclosure policies, and whether they may require reframing to address incompatibilities.

This study has several limitations. Our thematic findings cut across both HPE and general medical journal editor data; however, the latter sample is insufficient to fully portray disclosure expectations in the broader medical journal context. We focused on editors’ perceptions; the anonymized examples referenced in editor interviews were neither collected nor analyzed. Work is underway to analyze published disclosures to understand content and placement patterns. Our purposive sampling of experienced and AI-familiar journal editors excludes the perspectives of more AI-cautious editors and of peer reviewers, whose expectations and experiences will also influence the emerging norms of AI-use disclosure. In spite of these limitations, and the reality that a fluid AI environment renders findings quickly outdated, our study offers timely insights into the current ambiguities surrounding AI-use disclosure in HPE.

## Conclusion

“When in doubt, disclose”. Seems simple, but blurred thresholds of sufficiency and necessity complicate AI-use disclosure. Disclosure rules need to be explicit and dynamic. Attending to these thresholds and the factors blurring them may help HPE’s journals and authors navigate the shifting norms of AI-use disclosure.

## Supporting information

Supplemental Interview Guide

## Data Availability

Data produced in the present study is not available in order to protect participant confidentiality.

## Acknowledgements

We gratefully acknowledge the journal editors who participated in this study for sharing their experiences.

## Ethical approval

As indicated in the manuscript text, this study received institutional ethics approval from Western University’s Non-Medical Research Ethics Board (ID#125269).

