## Supplemental Interview Guide for "The blurred threshold of AI-use disclosure: International journal editors’ expectations of sufficiency and necessity"

**Interview guide for journal editors handling manuscripts that disclose AI-use**

**Introductory Script**

Thank you for taking the time to participate in this interview. My name is [Your Name], and I am conducting this research as part of a project aimed at understanding how GenAI use in manuscripts is disclosed and received by journals. Before we begin, I want to confirm that you have provided your consent to participate in this study.

The interview today will take approximately 30 minutes. During this time, we will discuss your experiences and perspectives related to generative AI use in manuscript submissions, as well as your journal’s policies and processes. Please remember that your participation is completely voluntary, and you are welcome to skip any questions you do not wish to answer or stop the interview at any time without any negative consequences.

Everything you share today will remain confidential. Your responses will be anonymized, and no identifying information will be included in any reports or publications arising from this research.

**Interview Questions**

1. What is your role in the editorial and peer review process at your journal?
2. What is your journal’s policy regarding the use of genAI in submitted manuscripts?
3. What does your journal require in terms of AI-use disclosure by authors?
   a. Where would an author find this information online?
4. Tell me about the last year or so at the journal: are you seeing AI-use disclosure in manuscripts submitted for peer review/sent out for peer review?
   a. If yes, what information are authors disclosing about how they used AI?
   b. If no, why do you think that is the case?
5. Please describe, referring to these submissions in an anonymous way, what a robust AI-use disclosure looks like? And an insufficient AI-use disclosure?
6. What is your sense, overall, of how peer-reviewers and associate editors are responding to manuscripts when AI use is disclosed?
7. What training or development has the journal provided for editors, associate editors and peer reviewers regarding handling of manuscripts disclosing AI use?
   a. Generally, what kinds of things are you doing in your journal related to [identifying, encouraging, discouraging, debating] the use of AI in submitted manuscripts?
8. Can you recall an instance where AI-use was a key part of the peer-review or editorial consideration of a manuscript? Please describe that case anonymously.
9. Is that the usual way AI-use comes up in editorial decision-making? If not, please share a different example to illustrate, in an anonymous manner.
10. Can you suggest other editorial roles at your journal that are centrally involved in the process of handling/wrestling with submissions that disclose AI-use? Do you think they might have insights relevant to our study?
